# Link between C-reactive protein-triglyceride glucose index and cardiovascular disease risk in diverse glycemic statuses: insights from the CHARLS

**DOI:** 10.1101/2025.11.14.25340283

**Authors:** Shifeng Qiu, Chen Yu, Siyun Guo, Xuewei Liu, Qiuxia Zhang, Hao Zheng, Kai Cui, Yuegang Wang, Guojun Chen, Yanmei Chen, Qingchun Zeng, Xue Li, Juefei Wu, Jiancheng Xiu

**Author notes:** Department of Cardiology, Nanfang Hospital, Southern Medical University, Guangzhou 510515, China. These authors have contributed equally to this work and share first authorship (Shifeng Qiu, Chen Yu, Siyun Guo). **Correspondence:** J.X. J.W. X.L.

## Abstract

**Objective:** The C-reactive protein-triglyceride-glucose index (CTI) is a novel biomarker that integrates measures of insulin resistance and inflammation. Its association with cardiovascular disease (CVD) risk across various glycemic statuses remains unclear.

**Methods:** This prospective study included 7,584 middle-aged and elderly participants from the China Health and Retirement Longitudinal Study (CHARLS). CTI was calculated using the formula 0.412*Ln(CRP) + Ln(TG × FPG)/2. Cox proportional hazards models and restricted cubic splines were employed to evaluate the relationship between CTI and CVD risk, with subgroup analyses conducted based on gender, age, and glycemic status..

**Results:** Over a median follow-up period of 108.1 months, 1,989 participants (26.23%) developed CVD. The incidence of CVD increased significantly across ascending CTI quartiles (Q1: 20.78% to Q4: 31.65%). A positive linear association was observed that remained consistent across both genders and age groups. Higher CTI quartiles (Q3/Q4) were associated with a significantly elevated risk of CVD, particularly among males (hazard ratio [HR] up to 1.58) and middle-aged individuals (HR 1.57). Notably, an elevated CTI was linked to an increased risk of CVD in participants with normal glucose regulation but not in those classified as prediabetic or diabetic.

**Conclusions:** The CTI serves as an independent predictor of CVD risk within the general middle-aged and elderly population. This association is most pronounced in individuals exhibiting normal glucose regulation, underscoring the potential utility of CTI for early risk stratification as well as the necessity for tailored prevention strategies informed by glycemic status.

## Introduction

Cardiovascular disease (CVD) is a leading cause of death and economic burden worldwide, especially in developing countries^[1–4]^. In China, it affects around 290 million people, causing approximately 3.7 million deaths annually^[5]^. CVD is the primary cause of death in both urban and rural China, with its prevalence increasing with age^[5]^, particularly among middle-aged and older adults^[6, 7]^. Despite progress in prevention, diagnosis, and treatment, CVD rates continue to rise^[8]^. Enhancing strategies to identify high-risk individuals and mitigate CVD risk is essential^[9]^.

Among the various risk factors for CVD, insulin resistance (IR) plays a crucial role in atherosclerotic cardiovascular disease^[10]^, making its assessment important for evaluating cardiovascular risk^[11]^. It contributes to CVD by causing inflammation, oxidative stress, and endothelial damage^[12–14]^, serving as both a cause and a poor prognosis indicator^[15–17]^. The triglyceride glucose (TyG) index is a common biomarker for IR^[18, 19]^ and is strongly linked to atherosclerosis and adverse cardiovascular outcomes^[20–22]^. C-reactive protein (CRP) is another key marker for CVD risk^[23, 24]^. The C-reactive protein-triglyceride glucose index (CTI)^[25]^, which reflects both IR and inflammation, is useful for predicting the risk of stroke^[26]^ and coronary heart disease (CHD) ^[27]^. Previous studies revealed a significant positive linear relationship between CTI and the occurrence of stroke^[26]^ and CHD^[27]^. However, its connection to CVD risk across different glycemic statuses remains unclear.

To fill this gap, we analyzed data from the China Health and Retirement Longitudinal Study (CHARLS) to explore the relationship between CTI and CVD risk under varying glucose metabolic conditions, aiming to enhance CTI’s practical application.

## Methods

### Study design

This analysis utilizes data from CHARLS, a national longitudinal survey that uses a multi-tiered stratified sampling approach among Chinese adults aged 45 years or older (http://charls.pku.edu.cn/). The inaugural national baseline survey (Wave 1) took place in 2011, amassing information from 17,708 middle-aged and elderly participants spanning 28 provinces, 150 regions, and 450 villages in China. Essential information from all participants was collected using standardized questionnaires, and follow-up visits will occur every 2–3 years to track their health status. For this analysis, we did not perform any interviews. So far, surveys have been carried out in five waves, specifically in 2011, 2013, 2015, 2018, and 2020, and we utilized the data from these waves. The detailed documentation of research methods and data collection procedures for implementing CHARLS is available in the literature^[28]^. Conducted in line with the Declaration of Helsinki, the CHARLS study was sanctioned by Peking University’s Institutional Review Board (IRB00001052-11015). Each participant provided written informed consent before engaging in the CHARLS study. The research adhered to the Strengthening the Reporting of Observational Studies in Epidemiology (STROBE) guidelines for reporting observational studies. Participants interviewed during 2011-2012 were used as the baseline for this study, with subsequent follow-ups in 2013, 2015, 2018 and 2020.

### Study population

The study’s participant screening process is shown in Fig. 1. Initially, 17,708 participants were identified in Wave 1. We excluded 10,124 participants due to various reasons: missing CRP, fasting plasma glucose (FPG), and triglycerides (TG) data (6,063 participants), missing stroke or heart disease history (125 participants in Wave 1 and 7 in Wave 5), loss to follow-up (2,494 participants), history of CVD (1,153 participants), age under 45 (220 participants), and missing hemoglobin A1c (HbA1c) data in Wave 5 (62 participants). This left 7,584 participants in the final study cohort.

**Fig. 1.**
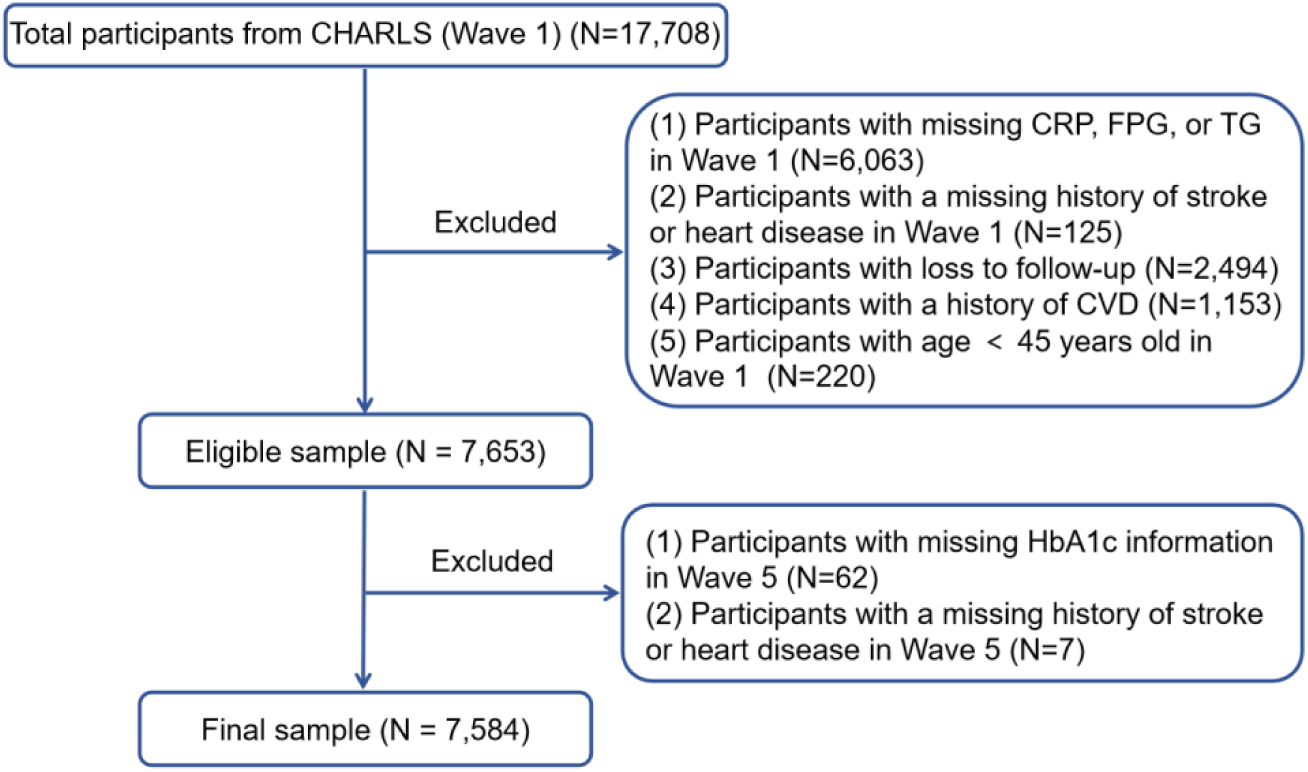
Flow chart of the study population.

### Exposure

Fasting venous blood samples were obtained by the Chinese Centre for Disease Control and Prevention’s medical team according to standard protocol and later examined at the central lab. Using an enzymatic colorimetric test, triglycerides and glucose were quantified. Within the assay, the coefficient of variation was 1.5% for triglycerides and 0.9% for glucose. The level of hsCRP was determined using an immunoturbidimetric method on a Hitachi 7180 chemistry analyzer (Hitachi, Tokyo, Japan). The coefficient of variation (CV) for measuring the blood marker was under 5%. The CTI index was calculated using the formula below^[29]^: CTI = 0.412 × Ln (CRP [mg/L]) + Ln (TG [mg/dl] × FPG [mg/dl])/2.

### Outcomes

The primary study outcome was the occurrence of CVD, encompassing coronary heart disease (CHD) and stroke. Consistent with prior studies^[30,31]^, the assessment of new cardiovascular events was conducted through these standardized questions: “Have you been told by a doctor that you have been diagnosed with a heart attack, coronary heart disease, angina, congestive heart failure, or other heart problems?” or “Have you been told by a doctor that you have been diagnosed with a stroke?” During the follow-up, participants reporting heart disease or stroke were recognized as having new cardiovascular disease. Trained interviewers evaluated the results using standardized questionnaires aligned with top international aging surveys. Diseases were classified using ICD-10 codes (International Statistical Classification of Diseases, 10th revision).

### Assessments of covariates

Trained interviewers carefully gathered the participants’ baseline data using structured questionnaires. (1) Demographic and lifestyle data: This section encompasses gender, age, residence, education level, marital status, and current smoking and drinking behaviors. (2) Body measurements: The measurements documented consist of height, weight, body mass index (BMI), as well as systolic and diastolic blood pressure (SBP and DBP). (3) Information on Disease and medication history: This includes information regarding existing conditions such as heart disease, hypertension, diabetes mellitus (DM), and dyslipidemia. (4) Laboratory test data: The laboratory data collected includes FPG, total cholesterol (TC), TG, high-density lipoprotein cholesterol (HDL-C), low-density lipoprotein cholesterol (LDL-C), HbA1c, leucocyte count (WBC), platelet count (PLT), hemoglobin (HGB), creatinine (CR), serum uric acid (SUA), triglyceride glucose (TyG) index, and TyG-BMI index.

### Definitions

Hypertension was characterized by any of these conditions: (1) SBP ≥ 140 mmHg, (2) DBP ≥ 90 mmHg, (3) self-reported hypertension diagnosed by a physician, (4) taking antihypertensive medications. DM was defined by meeting at least one of the following criteria: (1) FPG ≥ 126 mg/dL, or (2) HbA1c ≥ 6.5%, (3) and/or current use of antidiabetic medications, (4) and/or self-reported diabetes diagnosed by a doctor. Pre-DM was identified by an FPG of 100 to 125 mg/dL or an HbA1c of 5.7–6.4%. Normal glucose regulation (NGR) was defined as FPG < 100 mg/dL and HbA1c < 5.7%^[32]^. Dyslipidemia was recognized by TG ≥ 150 mg/dL, TC ≥ 240 mg/dL, HDL-C < 40 mg/dL, LDL-C ≥ 160 mg/dL, current use of lipid-lowering drugs, or a physician-confirmed diagnosis.

### Statistical analysis

Table S1 details the missing data in this study. Data are shown as means ± standard errors (means ± SEs) for normally distributed continuous variables, medians (interquartile ranges) for non-normal distributions, and percentages for categorical variables. To identify group differences, various tests were used: Welch’s *t*-test, ANOVA, Mann-Whitney for continuous variables, and chi-square for categorical variables. For ANOVA analyses, we performed post-hoc pairwise comparisons using the Tukey’s Honest Significant Difference (HSD) test to control for Type I error inflation due to multiple comparisons. For chi-squared tests, we performed pairwise comparisons with Bonferroni correction applied to the *p*-values.

Participants were divided into four groups based on CTI quartiles: Quartile (Q) 1 < 8.11, 8.11 ≤ Q2 < 8.61, 8.61 ≤ Q3 < 9.19, and Q4 ≥ 9.19. CTI was also analyzed as a continuous variable for robustness. Kaplan–Meier curves and log-rank tests assessed CVD incidence. Tolerance values and variance inflation factors (VIFs) were checked for collinearity between CTI and other covariates. The analysis showed that the VIFs for TC (VIF = 15.92), TG (VIF = 8.32), and LDL-C (VIF = 13.10) were above 5, leading to their exclusion from the multivariate model (Table S2). Cox regression models were used to examine the link between CTI and CVD incidence, with three models: Model 1 was unadjusted, Model 2 adjusted for gender, age, residence, marital status, education level, smoking status, and drinking status, and Model 3 contained additional adjustments for hypertension, dyslipidemia, BMI, SBP and DBP. The proportional hazards assumption of the Cox regression model was validated through the application of the Schoenfeld residuals test method. A fully adjusted restricted cubic splines (RCS) analysis explored the dose-response relationship between CTI and CVD risk. In cases of nonlinear relationships, potential threshold effects were identified by systematically testing all possible inflection points and selecting the most probable values. The receiver operating characteristics (ROC) curves assessed the predictive value of CTI, CRP, and TyG index for CVD incidence, with the area under the ROC curve (AUC) measuring CTI’s incremental effect. The study also evaluated CTI’s relationship with CVD risk across different genders, ages, and glycemic statuses (categorized as NGR, Pre-DM, and DM).

Subgroup analysis was prespecified based on previous literature^[26]^. Subgroup analyses assessed whether CTI’s effect on CVD incidence differed across demographic groups, stratified by gender, age (45–59 and ≥60 years), smoking and drinking status, hypertension, dyslipidemia, and BMI (< 24, 24 - 27.9, ≥28 kg/m²). To ensure robust findings, three sensitivity analyses were conducted: excluding non-fasting participants, performing multiple imputations for missing data, using logistic regression to examine CTI-CVD links, and calculating the E-value to determine the minimum association strength needed for an unmeasured confounder to negate observed associations. The E-value was calculated as E = RR + sqrt{RR × (RR−1)}^[33, 34]^. All statistical analyses utilized R software (version 4.3.3), with a two-sided *p* value < 0.05 regarded as statistically significant.

## Results

The study encompassed 7,584 participants from the CHARLS. The mean age of the participants was 57.72 ± 8.52 years, with 3,486 (45.97%) being male. Among the participants, 3,431 (45.24%) were classified as having NGR, 3,159 (41.65%) were identified as having Pre-DM, and 1,994 (13.11%) were diagnosed with DM. Furthermore, individuals in the higher quartiles of the CTI were more likely to reside in rural areas, possess a high school education or higher, and exhibit conditions such as hypertension, diabetes, and dyslipidemia compared to those in the lower quartiles of the CTI. Regarding anthropometric and laboratory assessments, elevated CTI quartiles were correlated with increased waist circumference, SBP, DBP, pulse rate, CRP, FPG, HbA1c, TC, TG, WBC, PLT, HGB, CR, SUA, TyG, and TyG-BMI. Conversely, higher CTI quartiles were associated with a higher likelihood of being unmarried, currently non-smoking, and having lower levels of HDL-C. The demographic and clinical characteristics of all participants are presented in Table 1. A comparison of baseline characteristics between included participants and those excluded from the analysis is provided in Table S3.

**Table 1.** Patient demographics and baseline characteristics.

| Variables | Total<br>(n = 7584) | Q1 (n = 1896) | Q2 (n = 1896) | Q3 (n = 1896) | Q4 (n = 1896) | p |
| --- | --- | --- | --- | --- | --- | --- |
| Gender, n(%) |  |  |  |  |  | **0.009** |
| Male | 3486 (45.97) | 914 (48.21) | 902 (47.57) | 834 (43.99) | 836 (44.09) |  |
| Female | 4098 (54.03) | 982 (51.79) | 994 (52.43) | 1062 (56.01) | 1060 (55.91) |  |
| Age, year | 57.72 ± 8.52 | 56.81 ± 8.56 | 57.81 ± 8.50 | 58.36 ± 8.58 | 57.90 ± 8.35 | **<0.001** |
| Residence, n(%) |  |  |  |  |  | **<0.001** |
| Rural | 5071 (66.86) | 1357 (71.57) | 1332 (70.25) | 1217 (64.19) | 1165 (61.45) |  |
| Urban | 2513 (33.14) | 539 (28.43) | 564 (29.75) | 679 (35.81) | 731 (38.55) |  |
| Marital status, n(%) |  |  |  |  |  | 0.774 |
| Married | 724 (9.55) | 181 (9.55) | 187 (9.86) | 186 (9.81) | 170 (8.97) |  |
| Other | 6860 (90.45) | 1715 (90.45) | 1709 (90.14) | 1710 (90.19) | 1726 (91.03) |  |
| Education level, n(%) |  |  |  |  |  | 0.446 |
| No formal education | 3536 (46.68) | 884 (46.67) | 892 (47.10) | 908 (47.99) | 852 (44.96) |  |
| Primary school | 1622 (21.41) | 394 (20.80) | 423 (22.33) | 401 (21.19) | 404 (21.32) |  |
| Middle school | 1602 (21.15) | 421 (22.23) | 375 (19.80) | 386 (20.40) | 420 (22.16) |  |
| High school or above | 815 (10.76) | 195 (10.30) | 204 (10.77) | 197 (10.41) | 219 (11.56) |  |
| Smoking, n(%) | 2223 (29.83) | 587 (31.49) | 583 (31.31) | 529 (28.43) | 524 (28.11) | **0.032** |
| Drinking, n(%) | 2596 (34.24) | 704 (37.15) | 674 (35.57) | 600 (31.65) | 618 (32.59) | **<0.001** |
| BMI, kg/m<sup>2</sup> | 24.16 ± 32.46 | 22.04 ± 3.08 | 23.78 ± 20.75 | 25.77 ± 61.03 | 25.06 ± 3.87 | **0.006** |
| Waist, cm | 83.84 ± 12.43 | 79.68 ± 10.58 | 82.19 ± 11.72 | 85.18 ± 12.65 | 88.39 ± 12.94 | **<0.001** |
| SBP, mmHg | 127.85 ± 20.54 | 123.25 ± 19.83 | 125.79 ± 19.80 | 129.62 ± 20.47 | 132.82 ± 20.77 | **<0.001** |
| DBP, mmHg | 74.90 ± 11.97 | 72.54 ± 11.93 | 73.74 ± 11.54 | 75.69 ± 11.81 | 77.71 ± 11.97 | **<0.001** |
| Pulse, bpm | 71.90 ± 10.16 | 70.39 ± 9.89 | 71.32 ± 9.99 | 71.93 ± 9.99 | 73.99 ± 10.45 | **<0.001** |
| Complication |  |  |  |  |  |  |
| Hypertension, n(%) | 1654 (21.92) | 266 (14.08) | 325 (17.25) | 463 (24.45) | 600 (31.93) | **<0.001** |
| Diabetes, n(%) | 355 (4.72) | 34 (1.80) | 43 (2.29) | 87 (4.63) | 191 (10.18) | **<0.001** |
| Dyslipidemia, n(%) | 587 (7.89) | 67 (3.58) | 105 (5.65) | 142 (7.65) | 273 (14.72) | **<0.001** |
| Cancer, n(%) | 51 (0.67) | 10 (0.53) | 13 (0.69) | 15 (0.79) | 13 (0.69) | 0.800 |
| Laboratory indicators |  |  |  |  |  |  |
| CRP, mg/dl | 0.95 (0.52, 1.96) | 0.47 (0.33,0.70) | 0.76 (0.50,1.26) | 1.28 (0.75,2.28) | 2.32 (1.25,4.82) | **<0.001** |
| FPG, mg/dl | 102.06 (94.32, 112.50) | 96.12 (89.28,103.32) | 99.54 (93.24,107.10) | 103.14 (95.94,111.78) | 113.40 (102.42,135.18) | **<0.001** |
| HbA1c, % | 5.10 (4.90, 5.40) | 5.00 (4.80,5.30) | 5.10 (4.88,5.40) | 5.10 (4.90,5.40) | 5.30 (5.00,5.70) | **<0.001** |
| GMS, n(%) |  |  |  |  |  | **<0.001** |
| NGR | 3431 (45.24) | 1186 (62.55) | 985 (51.95) | 780 (41.14) | 480 (25.32) |  |
| Pre-DM | 3159 (41.65) | 636 (33.54) | 779 (41.09) | 900 (47.47) | 844 (44.51) |  |
| DM | 994 (13.11) | 74 (3.90) | 132 (6.96) | 216 (11.39) | 572 (30.17) |  |
| TC, mg/dl | 190.59<br>(167.40,<br>214.95) | 180.93<br>(160.44,202.96) | 188.27<br>(166.62,211.08) | 193.30<br>(170.10,217.27) | 200.65<br>(174.36,230.12) | **<0.001** |
| TG, mg/dl | 104.43 (74.34,<br>153.99) | 65.49<br>(53.10,79.65) | 94.69<br>(77.00,113.28) | 125.23<br>(98.90,153.99) | 196.47<br>(143.37,273.46) | **<0.001** |
| HDL-C,<br>mg/dl | 49.48 (40.59,<br>59.92) | 59.15<br>(50.26,69.20) | 52.58<br>(45.23,61.47) | 47.55<br>(40.21,56.44) | 39.43<br>(32.86,47.94) | **<0.001** |
| LDL-C,<br>mg/dl | 114.43 (93.56,<br>136.86) | 109.79<br>(90.85,128.74) | 116.75<br>(96.65,138.02) | 119.85<br>(98.58,142.66) | 112.50<br>(86.60,139.95) | **<0.001** |
| WBC,<br>10<sup>9</sup>/L | 5.90 (4.91,<br>7.20) | 5.50 (4.60,6.50) | 5.76 (4.77,6.80) | 6.10 (5.10,7.40) | 6.41 (5.40,7.80) | **<0.001** |
| PLT, 10<sup>9</sup>/L | 207.00<br>(163.00,<br>254.00) | 203.00<br>(161.50,247.00) | 204.00<br>(159.00,252.00) | 209.50<br>(164.00,260.00) | 213.00<br>(167.00,261.00) | **<0.001** |
| HGB, g/dl | 14.20 (13.00,<br>15.50) | 14.00<br>(12.80,15.30) | 14.20<br>(13.00,15.50) | 14.30<br>(13.10,15.50) | 14.50<br>(13.20,15.80) | **<0.001** |
| CR, mg/dl | 0.75 (0.64,<br>0.87) | 0.75 (0.63,0.85) | 0.75 (0.64,0.87) | 0.76 (0.64,0.88) | 0.76 (0.66,0.89) | **<0.001** |
| SUA, mg/dl | 4.25 (3.55,<br>5.10) | 3.98 (3.37,4.74) | 4.13 (3.47,4.97) | 4.34 (3.65,5.20) | 4.62 (3.80,5.50) | **<0.001** |
| TyG | 8.58 (8.22,<br>9.03) | 8.06 (7.86,8.25) | 8.47 (8.27,8.66) | 8.79 (8.54,9.01) | 9.36 (9.01,9.77) | **<0.001** |
| TyG-BMI | 198.54<br>(175.68,<br>228.19) | 175.05<br>(159.31,191.79) | 189.75<br>(173.06,209.37) | 209.34<br>(187.60,230.94) | 235.00<br>(207.98,262.05) | **<0.001** |
| CVD, n(%) | 1989 (26.23) | 394 (20.78) | 474 (25.00) | 521 (27.48) | 600 (31.65) | **<0.001** |

Abbreviation: BMI - body mass index, SBP - systolic blood pressure, DBP - diastolic blood
pressure, CRP - C-reactive protein, FPG - fasting plasma glucose, HbA1c - hemoglobin A1c,
GMS - glycemic statuses, NGR - Normal glucose regulation, Pre-DM - prediabetes mellitus, DM -
diabetes mellitus, TC - total cholesterol, TG - triglycerides, HDL-C - high-density lipoprotein
cholesterol, LDL-C - low-density lipoprotein cholesterol, WBC - leucocyte count, PLT - platelet
count, HGB - hemoglobin, CR - creatinine, SUA - serum uric acid, TyG - triglyceride glucose.

### Association between the CTI and the risk of CVD

During a median follow-up of 108.10 months (IQR 106.09–108.10), 1,989 (26.23%) participants developed CVD, with incidence rising from 20.78% in Q1 to 31.65% in Q4. The Kaplan–Meier curve showed a significant increase in CVD events across quartiles (log-rank test P < 0.001, Fig. 2A). Cox regression indicated a 6% higher CVD risk per 1-unit CTI increase (HR 1.06, 95% CI 1.01, 1.11), but no significant risk increase for CRP and TyG index (Table 2) (Table S4). Compared to Q1, CVD risk rose with higher CTI levels: hazard ratios (HRs) were 1.25 (95% CI 1.08, 1.44) for Q3, and 1.29 (95% CI 1.11, 1.49) for Q4, but no significance in Q2 (*p* > 0.008). RCS analysis confirmed a significant linear relationship between CTI and CVD events (Fig. 3A). The highest quartile HR were 1.23 (95% CI 1.07, 1.42) for CRP and 1.14 (95% CI 0.99, 1.32) for the TyG index, both lower than CTI’s HR of 1.29 (95% CI 1.11, 1.49) (Table S4, Table 2). ROC curves showed CTI had the highest diagnostic efficacy for CVD incidence (AUC 0.557, 95% CI 0.543, 0.572), followed by CRP (AUC 0.546, 95% CI 0.532, 0.561) and the TyG index (AUC 0.544, 95% CI 0.529, 0.559) (Fig. S1). This suggests CTI is a more effective CVD risk predictor than CRP and the TyG index.

**Fig. 2.**
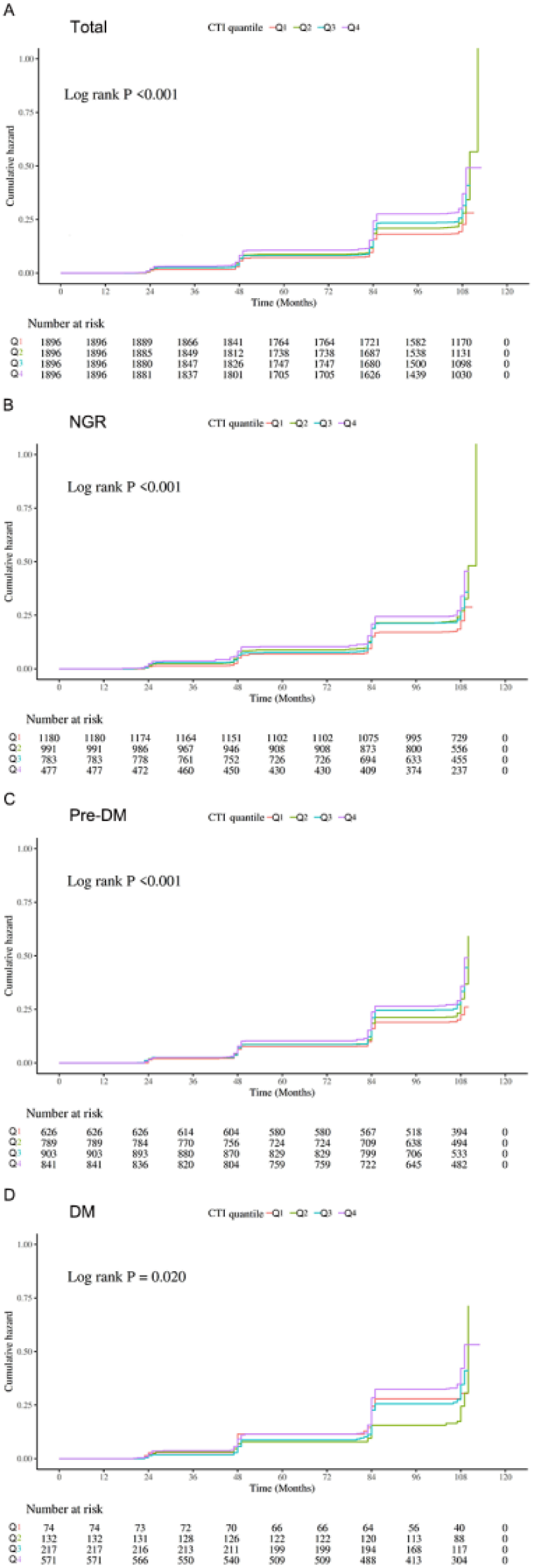
The Kaplan–Meier curves analysis depict the cumulative incidence of CVD across the CTI index quartiles. (A) total participants, (B) participants with NGR, (C) participants with Pre-DM, (D) participants with DM.

**Table 2.**
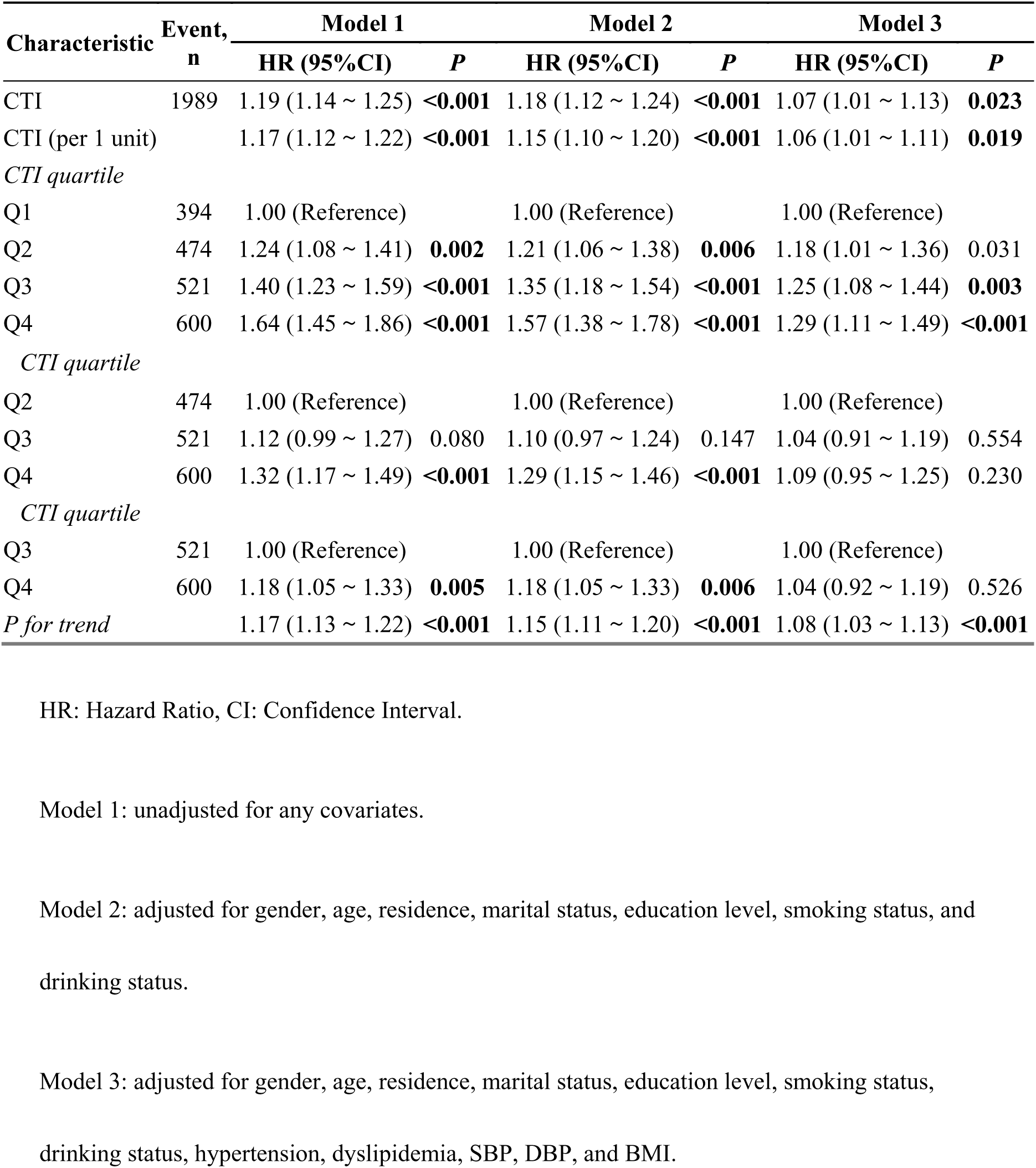
Association between the CTI and CVD incidence.

**Fig. 3.**
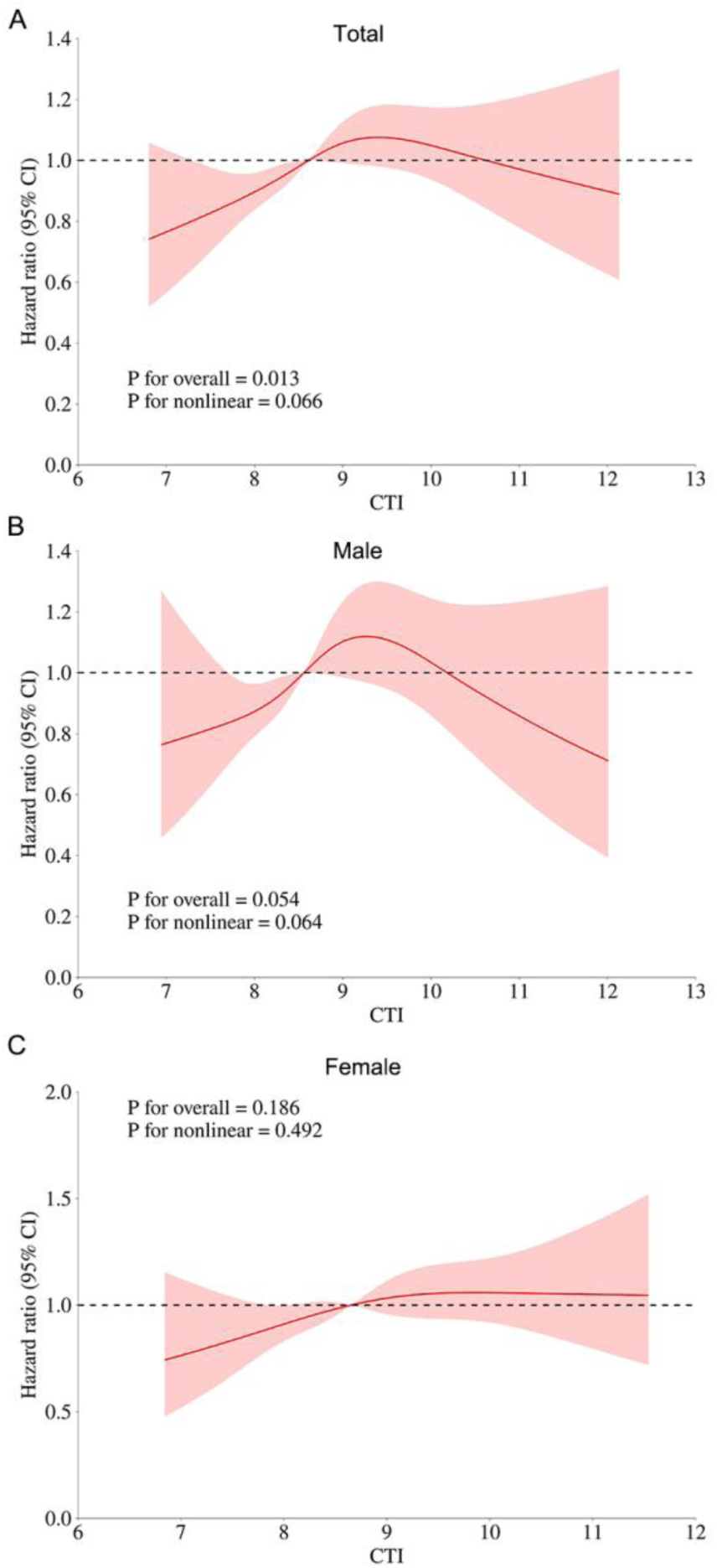
Association of the CTI index and the risk of CVD according to gender. (A) total participants, (B) male, (C) female. Adjusted for gender, age, residence, marital status, education level, smoking status, drinking status, hypertension, dyslipidemia, SBP, DBP, and BMI.

This relationship was significant across genders after adjusting for variables, with similar HRs for males and females. Higher CTI levels correlated with increased CVD risk, with significant HRs in both genders. Compared to Q1, CVD risk increased with higher CTI levels, with HRs of 1.39 (95% CI 1.14, 1.70), and 1.58 (95% CI 1.29, 1.93) for Q3, and Q4 in males, and with HRs of 1.56 (95% CI 1.32, 1.86) for Q4 in females (P for trend < 0.05) in model 2. When CTI was used as a quantitative variable, it was not statistically significant between CTI and CVD for males and females (Fig. 3B, and 3C). No interaction was found between CTI and CVD across gender groups (P for interaction = 0.604).

**Table 3.** Association between the CTI and CVD incidence according to gender.

| Gender | Characteristic | Event,<br>n | Model 1 |  | Model 2 |  | Model 3 |  |
| --- | --- | --- | --- | --- | --- | --- | --- | --- |
|  |  |  | HR (95%CI) | p | HR (95%CI) | p | HR (95%CI) | p |
| Male | CTI | 846 | 1.17 (1.09 ~ 1.26) | <0.001 | 1.10 (1.01 ~ 1.20) | **0.021** | 1.07 (0.97 ~ 1.17) | 0.160 |
|  | CTI (per 1 unit) |  | 1.14 (1.07 ~ 1.22) | <0.001 | 1.17 (1.10 ~ 1.25) | <0.001 | 1.06 (0.98 ~ 1.14) | 0.160 |
|  | *CTI quartile* |  |  |  |  |  |  |  |
|  | Q1 | 181 | 1.00 (Reference) |  | 1.00 (Reference) |  | 1.00 (Reference) |  |
|  | Q2 | 208 | 1.17 (0.96 ~ 1.42) | 0.127 | 1.17 (0.96 ~ 1.44) | 0.122 | 1.18 (0.94 ~ 1.48) | 0.150 |
|  | Q3 | 219 | 1.36 (1.11 ~ 1.65) | **0.002** | 1.39 (1.14 ~ 1.70) | **0.001** | 1.28 (1.03 ~ 1.61) | 0.028 |
|  | Q4 | 238 | 1.52 (1.25 ~ 1.84) | <0.001 | 1.58 (1.29 ~ 1.93) | <0.001 | 1.29 (1.03 ~ 1.62) | 0.026 |
|  | *CTI quartile* |  |  |  |  |  |  |  |
|  | Q2 | 208 | 1.00 (Reference) |  | 1.00 (Reference) |  | 1.00 (Reference) |  |
|  | Q3 | 219 | 1.09 (0.90 ~ 1.32) | 0.359 | 1.11 (0.92 ~ 1.35) | 0.289 | 1.04 (0.84 ~ 1.28) | 0.748 |
|  | Q4 | 238 | 1.26 (1.05 ~ 1.52) | 0.013 | 1.32 (1.09 ~ 1.59) | **0.004** | 1.10 (0.88 ~ 1.36) | 0.406 |
|  | *CTI quartile* |  |  |  |  |  |  |  |
|  | Q3 | 219 | 1.00 (Reference) |  | 1.00 (Reference) |  | 1.00 (Reference) |  |
|  | Q4 | 238 | 1.16 (0.96 ~ 1.38) | 0.116 | 1.19 (0.99 ~ 1.43) | 0.070 | 1.06 (0.86 ~ 1.30) | 0.588 |
|  | *P for trend* |  | 1.15 (1.08 ~ 1.22) | <0.001 | 1.17 (1.10 ~ 1.24) | <0.001 | 1.09 (1.01 ~ 1.17) | 0.021 |
| Female | CTI | 1143 | 1.21 (1.13 ~ 1.29) | <0.001 | 1.08 (1.01 ~ 1.15) | 0.042 | 1.07 (0.99 ~ 1.16) | 0.069 |
|  | CTI (per 1 unit) |  | 1.17 (1.11 ~ 1.24) | <0.001 | 1.13 (1.07 ~ 1.20) | <0.001 | 1.06 (1.00 ~ 1.13) | 0.069 |
|  | *CTI quartile* |  |  |  |  |  |  |  |
|  | Q1 | 209 | 1.00 (Reference) |  | 1.00 (Reference) |  | 1.00 (Reference) |  |
|  | Q2 | 270 | 1.28 (1.07 ~ 1.54) | **0.007** | 1.23 (1.03 ~ 1.48) | 0.023 | 1.18 (0.97 ~ 1.44) | 0.092 |
|  | Q3 | 302 | 1.37 (1.14 ~ 1.63) | <0.001 | 1.27 (1.06 ~ 1.51) | 0.009 | 1.18 (0.98 ~ 1.44) | 0.087 |
|  | Q4 | 362 | 1.71 (1.44 ~ 2.03) | <0.001 | 1.56 (1.32 ~ 1.86) | <0.001 | 1.29 (1.06 ~ 1.56) | 0.010 |
|  | *CTI quartile* |  |  |  |  |  |  |  |
|  | Q2 | 270 | 1.00 (Reference) |  | 1.00 (Reference) |  | 1.00 (Reference) |  |
|  | Q3 | 302 | 1.12 (0.95 ~ 1.32) | 0.181 | 1.09 (0.93 ~ 1.29) | 0.290 | 1.05 (0.88 ~ 1.26) | 0.571 |
|  | Q4 | 362 | 1.36 (1.16 ~ 1.59) | <0.001 | 1.30 (1.11 ~ 1.53) | **0.001** | 1.12 (0.94 ~ 1.33) | 0.220 |

| Gender Characteristic | Event,<br>n | Model 1 |  | Model 2 |  | Model 3 |  |
| --- | --- | --- | --- | --- | --- | --- | --- |
|  |  | HR (95%CI) | *p* | HR (95%CI) | *p* | HR (95%CI) | *p* |
| *CTI quartile* |  |  |  |  |  |  |  |
| Q3 | 302 | 1.00 (Reference) |  | 1.00 (Reference) |  | 1.00 (Reference) |  |
| Q4 | 362 | 1.21 (1.04 ~ 1.41) | 0.015 | 1.19 (1.02 ~ 1.39) | 0.028 | 1.06 (0.90 ~ 1.26) | 0.497 |
| *P for trend* |  | 1.18 (1.12 ~ 1.24) | **<0.001** | 1.15 (1.09 ~ 1.21) | **<0.001** | 1.08 (1.01 ~ 1.14) | **0.016** |

HR: Hazard Ratio, CI: Confidence Interval.
Model 1: unadjusted for any covariates.
Model 2: adjusted for age, residence, marital status, education level, smoking status, and drinking status.
Model 3: adjusted for age, residence, marital status, education level, smoking status, drinking status, hypertension, dyslipidemia, SBP, DBP, and BMI.

Table 4 indicates a notable link between CTI and CVD risk across different age groups after adjustments, with elevated CTI levels associated with a higher risk of CVD. Middle-aged participants (45–59 years) had a higher HR (1.57, 95% CI 1.15–2.15) in model 3 than elderly participants (≥ 60 years) with an HR of 1.48 (95% CI 0.22–1.80) in model 2. No interaction was found between CTI and CVD by age group (P for interaction = 0.414), and the association was similar for both middle-aged and elderly participants. Whereas, RCS analysis showed no clear relationship between CTI and CVD events in the middle-aged and elderly (Fig. 4).

**Table 4.**
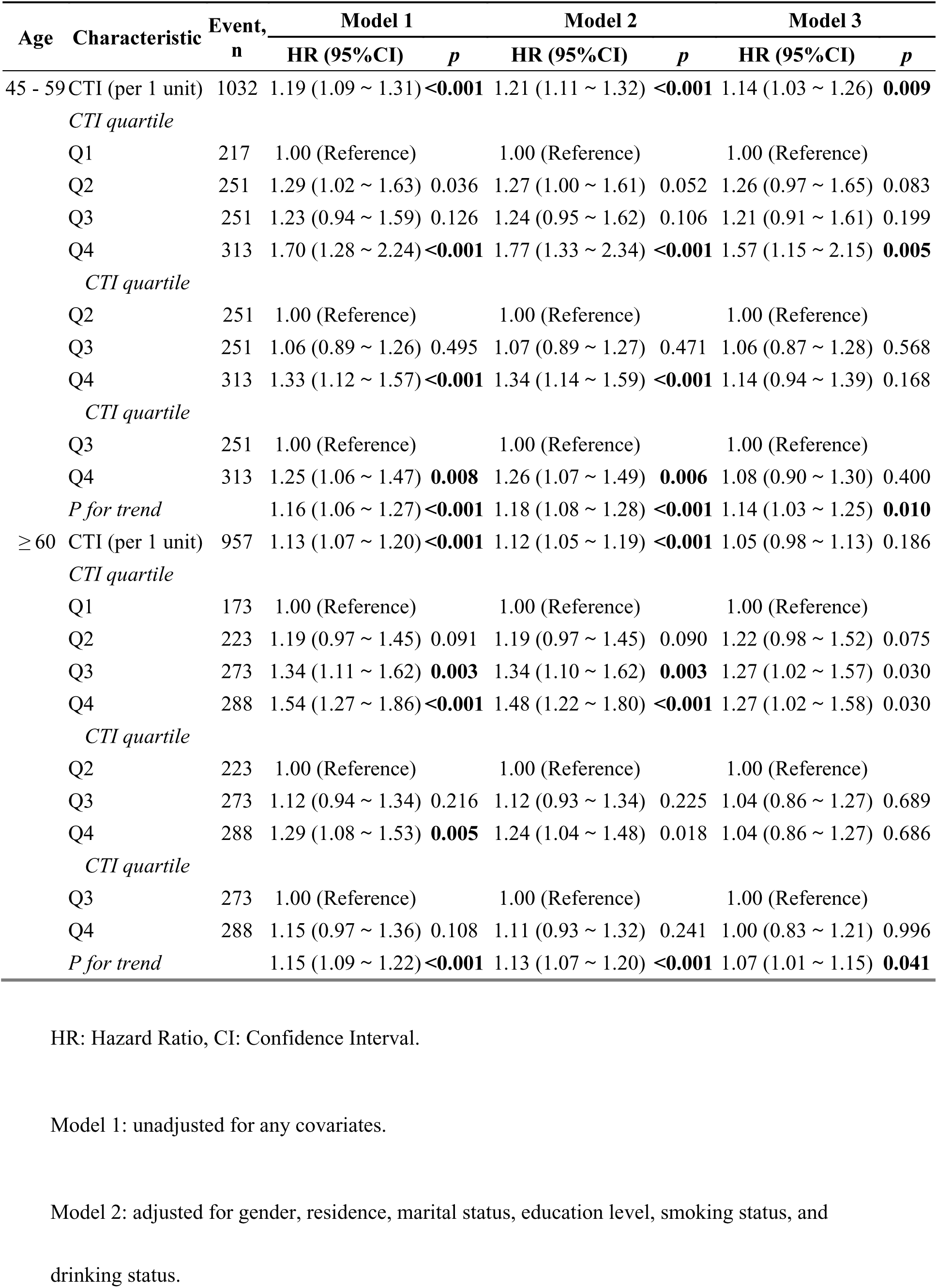
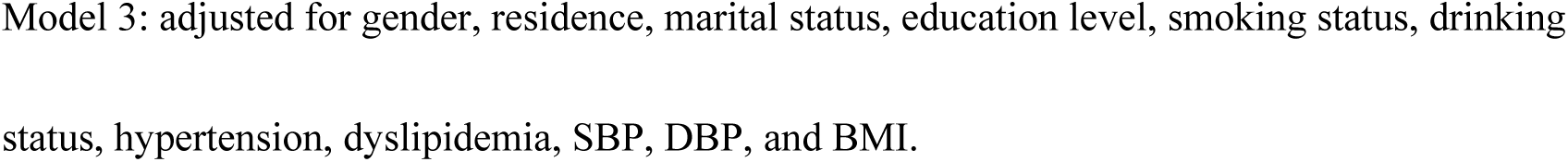
Association between the CTI and CVD incidence according to age.

**Fig. 4.**
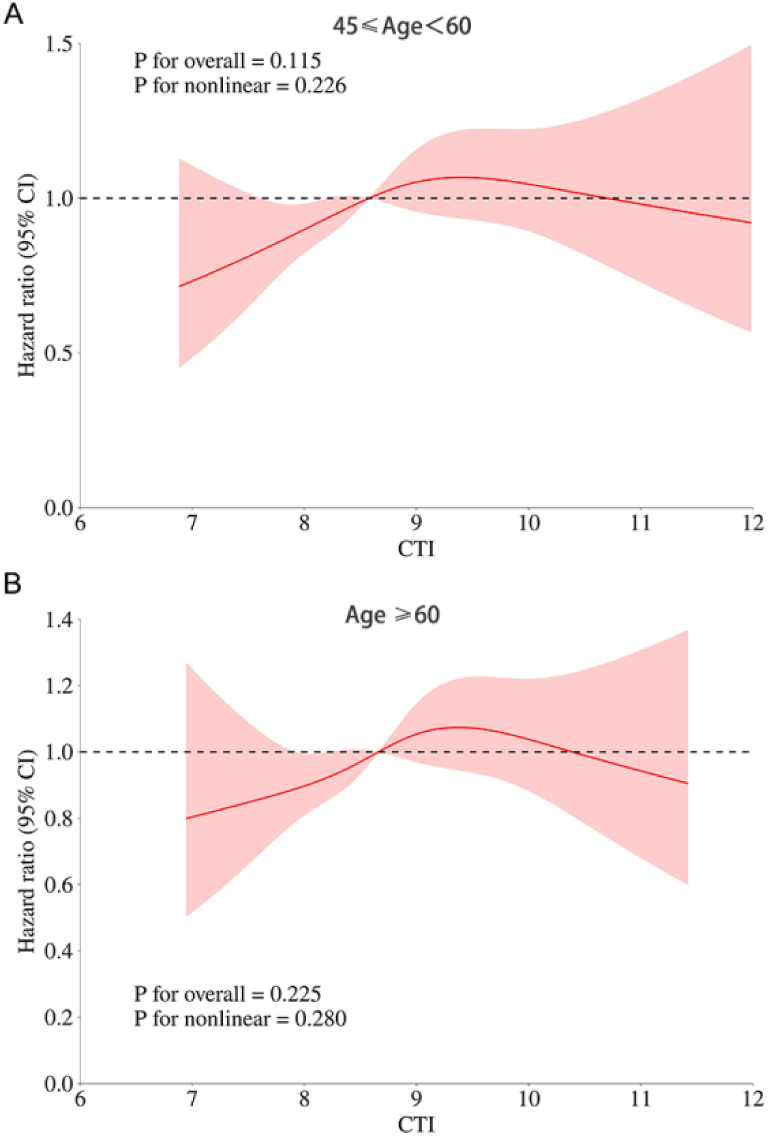
Association of the CTI index and the risk of CVD according to age. (A) 45 ≤ age < 60, (B) age ≥ 60. Adjusted for gender, residence, marital status, education level, smoking status, drinking status, hypertension, dyslipidemia, SBP, DBP, and BMI.

### Association between the CTI and CVD risk according to glycemic status

During follow-up, 813 (40.8%) with NGR, 865 (43.5%) with Pre-DM, and 311 (15.6%) with DM experienced their first CVD. Kaplan–Meier curves highlighted significant differences in CVD incidence among NGR, Pre-DM, and DM across CTI groups (all log-rank test *p* < 0.05, Fig. 2B, C, and D). Table 5 shows a significant association between CTI and CVD risk in NGR and Pre-DM when CTI served as a classification variable. However, individuals with DM showed no significant differences (all *p* > 0.008). Specifically, a 1 unit increase in CTI was linked to a 9% higher risk of CVD in people with NGR (HR 1.09, 95% CI 1.01, 1.17), while no notable difference was observed in those with Pre-DM and DM (all *p* > 0.05). Additionally, the RCS analysis failed to identify a significant dose–response link between the CTI and CVD occurrences in individuals with different glycemic statuses (Fig. 5A, B, and C).

**Table 5.**
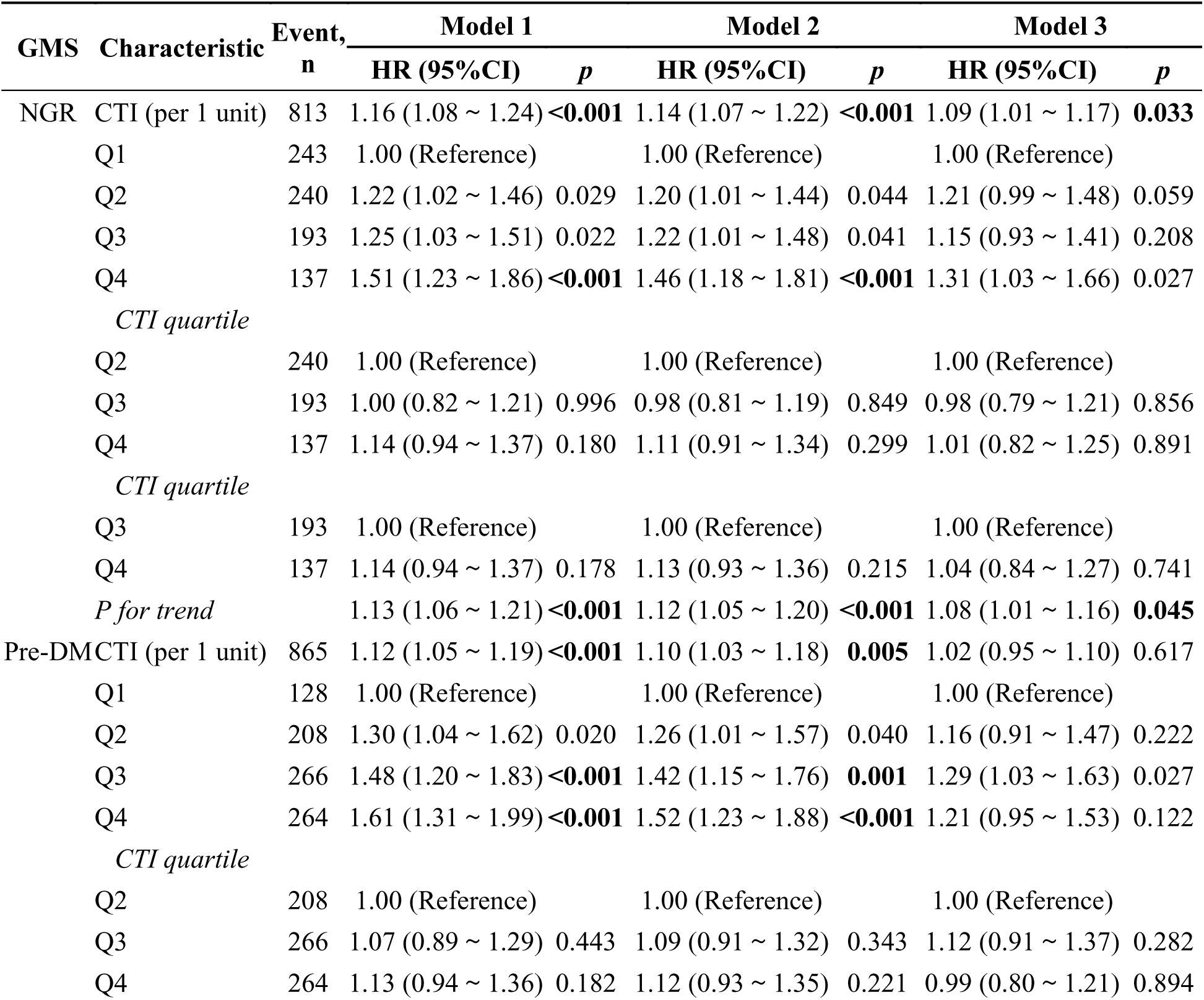
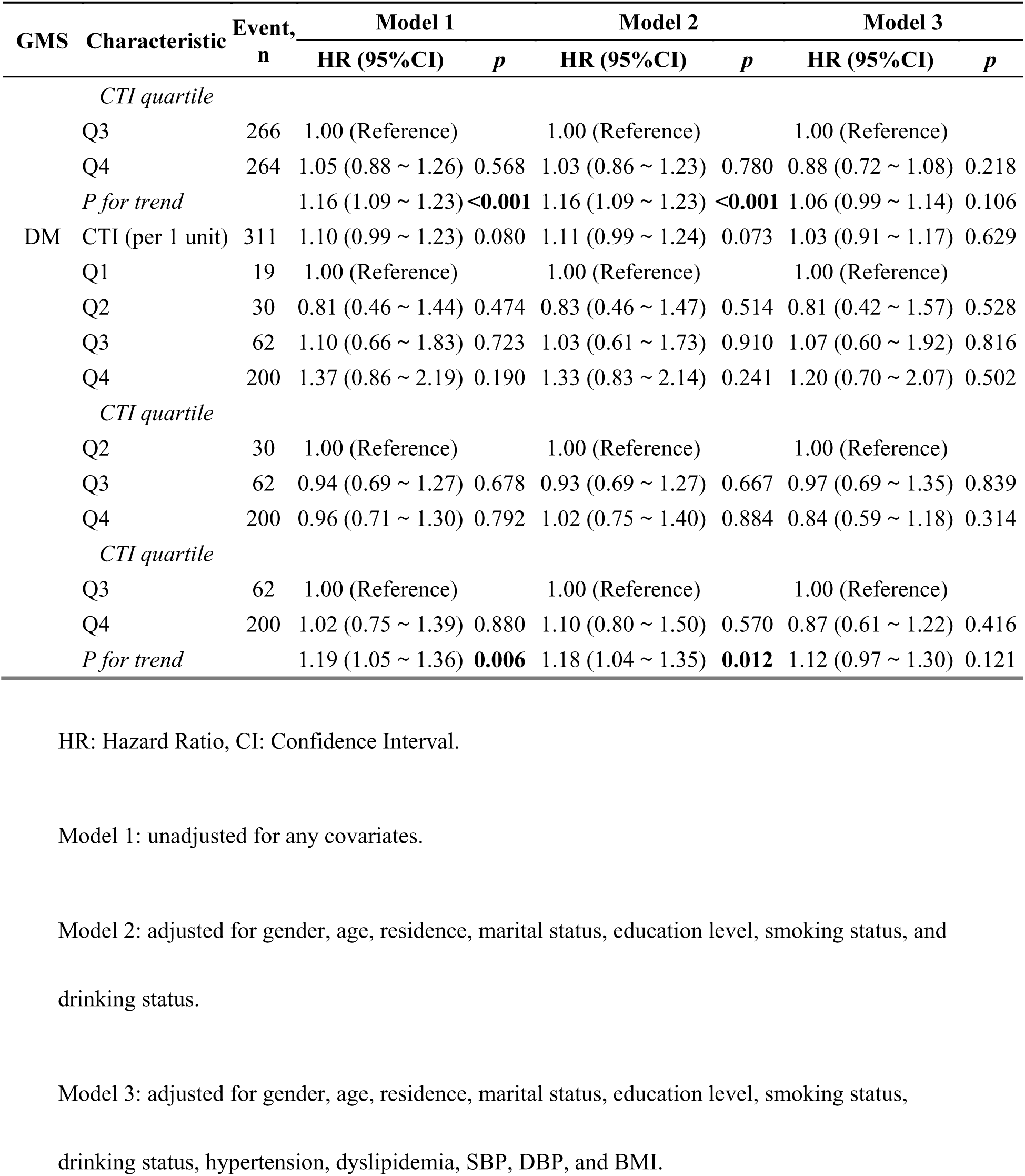
Association between the CTI and CVD incidence according to glucose regulation state.

**Fig. 5.**
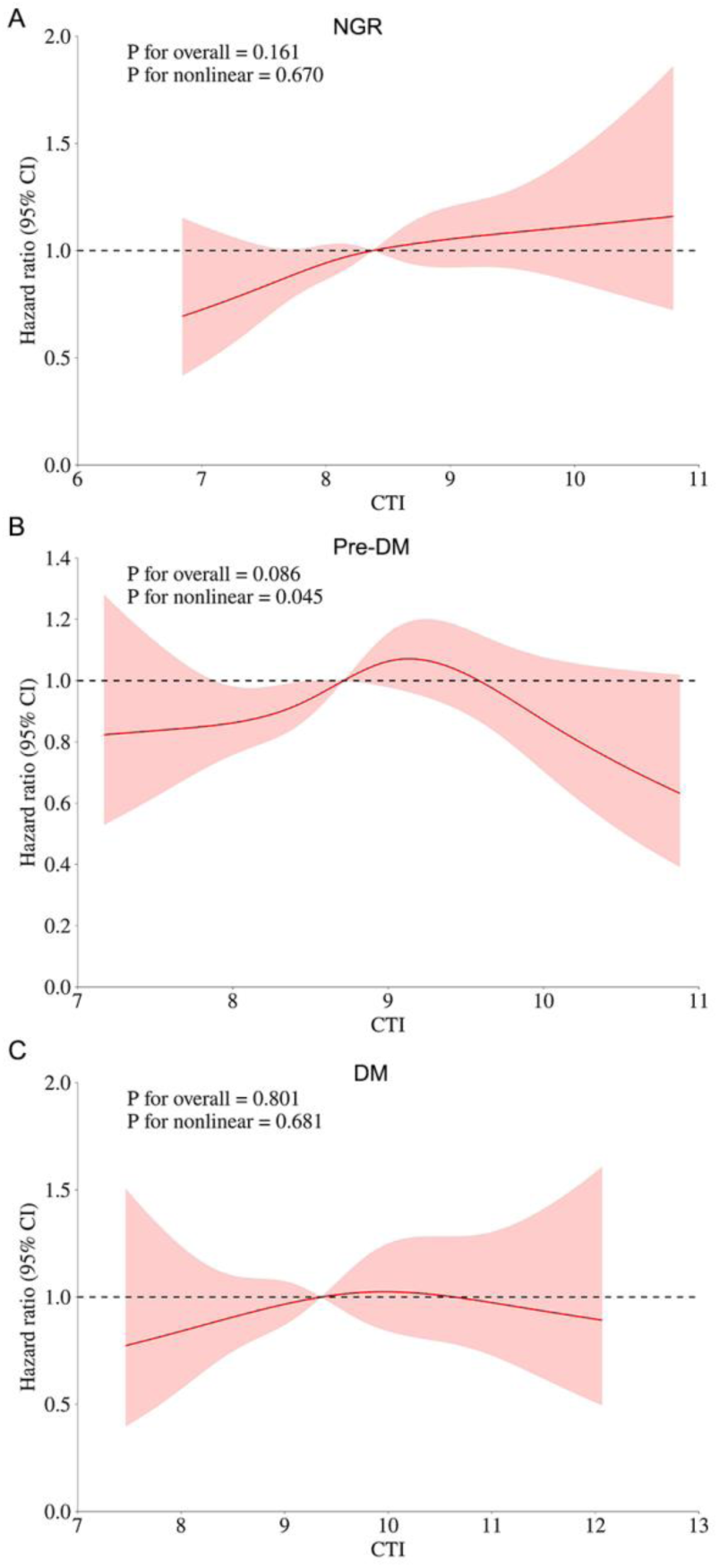
Association of the CTI index and the risk of CVD according to glucose metabolic states. (A) participants with NGR, (B) participants with Pre-DM, (C) participants with DM. Adjusted for gender, age, residence, marital status, education level, smoking status, drinking status, hypertension, dyslipidemia, dyslipidemia medications, SBP, DBP, and BMI.

As a continuous variable, CTI showed no significant difference across NGR, Pre-DM, and DM groups in both males and females. As a classified variable, CVD risk was significantly higher in Q4 for Pre-DM (HR 1.88) and in Q3 for NGR (HR 1.51), compared to Q1 in males. Among female participants, the risk of CVD was notably greater in Q4 compared to Q1 for those with NGR (HR 1.62), with similar risks in Q3 for Pre-DM (HR 1.49), but not for individuals with DM. (Table 6)

**Table 6.** Association between the CTI and CVD incidence according to gender and glucose regulation state.

| Gender | GMS | Characteristic | Event,<br>n | Model 1 |  | Model 2 |  | Model 3 |  |
| --- | --- | --- | --- | --- | --- | --- | --- | --- | --- |
|  |  |  |  | HR (95%CI) | *p* | HR (95%CI) | *p* | HR (95%CI) | *p* |
| Male | NGR | CTI (per 1 unit) | 356 | 1.10 (1.00 ~ 1.22) | 0.057 | 1.13 (1.02 ~ 1.26) | **0.019** | 1.07 (0.95 ~ 1.20) | 0.250 |
|  |  | Q1 | 114 | 1.00 (Reference) |  | 1.00 (Reference) |  | 1.00 (Reference) |  |
|  |  | Q2 | 100 | 1.07 (0.82 ~ 1.40) | 0.607 | 1.09 (0.83 ~ 1.44) | 0.548 | 1.06 (0.78 ~ 1.44) | 0.708 |
|  |  | Q3 | 95 | 1.40 (1.07 ~ 1.84) | **0.015** | 1.51 (1.14 ~ 2.00) | **0.004** | 1.44 (1.06 ~ 1.96) | 0.021 |
|  |  | Q4 | 47 | 1.12 (0.80 ~ 1.58) | 0.507 | 1.18 (0.84 ~ 1.68) | 0.342 | 1.10 (0.75 ~ 1.62) | 0.619 |
|  |  | *P for trend* |  | 1.09 (0.99 ~ 1.20) | 0.096 | 1.11 (1.01 ~ 1.23) | **0.037** | 1.09 (0.97 ~ 1.22) | 0.150 |
|  | Pre-DM | CTI (per 1 unit) | 365 | 1.20 (1.09 ~ 1.32) | **<0.001** | 1.21 (1.09 ~ 1.35) | **<0.001** | 1.09 (0.96 ~ 1.23) | 0.167 |
|  |  | Q1 | 60 | 1.00 (Reference) |  | 1.00 (Reference) |  | 1.00 (Reference) |  |
|  |  | Q2 | 92 | 1.33 (0.96 ~ 1.84) | 0.089 | 1.28 (0.92 ~ 1.79) | 0.137 | 1.31 (0.92 ~ 1.88) | 0.138 |
|  |  | Q3 | 96 | 1.32 (0.96 ~ 1.82) | 0.092 | 1.27 (0.92 ~ 1.77) | 0.151 | 1.14 (0.79 ~ 1.65) | 0.491 |
|  |  | Q4 | 117 | 1.91 (1.40 ~ 2.60) | **<0.001** | 1.88 (1.37 ~ 2.58) | **<0.001** | 1.46 (1.01 ~ 2.11) | 0.042 |
|  |  | *P for trend* |  | 1.22 (1.10 ~ 1.34) | **<0.001** | 1.21 (1.10 ~ 1.34) | **<0.001** | 1.10 (0.98 ~ 1.24) | 0.093 |
|  | DM | CTI (per 1 unit) | 125 | 1.05 (0.88 ~ 1.25) | 0.575 | 1.07 (0.88 ~ 1.29) | 0.497 | 0.86 (0.68 ~ 1.08) | 0.201 |
|  |  | Q1 | 7 | 1.00 (Reference) |  | 1.00 (Reference) |  | 1.00 (Reference) |  |
|  |  | Q2 | 16 | 1.11 (0.46 ~ 2.70) | 0.820 | 1.26 (0.51 ~ 3.08) | 0.615 | 1.25 (0.46 ~ 3.37) | 0.659 |
|  |  | Q3 | 28 | 1.47 (0.64 ~ 3.36) | 0.362 | 1.64 (0.71 ~ 3.82) | 0.248 | 1.38 (0.55 ~ 3.47) | 0.499 |
|  |  | Q4 | 74 | 1.42 (0.65 ~ 3.08) | 0.375 | 1.57 (0.71 ~ 3.49) | 0.266 | 1.14 (0.47 ~ 2.78) | 0.772 |
|  |  | *P for trend* |  | 1.11 (0.92 ~ 1.34) | 0.272 | 1.13 (0.93 ~ 1.38) | 0.231 | 0.99 (0.79 ~ 1.25) | 0.963 |
| Female | NGR | CTI (per 1 unit) | 457 | 1.20 (1.10 ~ 1.31) | **<0.001** | 1.14 (1.04 ~ 1.25) | **0.004** | 1.10 (0.99 ~ 1.21) | 0.079 |
|  |  | Q1 | 129 | 1.00 (Reference) |  | 1.00 (Reference) |  | 1.00 (Reference) |  |
|  |  | Q2 | 140 | 1.35 (1.07 ~ 1.72) | 0.013 | 1.29 (1.01 ~ 1.64) | 0.041 | 1.36 (1.04 ~ 1.76) | 0.023 |
|  |  | Q3 | 98 | 1.13 (0.87 ~ 1.47) | 0.364 | 1.02 (0.78 ~ 1.33) | 0.865 | 0.96 (0.72 ~ 1.29) | 0.803 |
|  |  | Q4 | 90 | 1.84 (1.41 ~ 2.41) | **<0.001** | 1.62 (1.23 ~ 2.13) | **<0.001** | 1.45 (1.07 ~ 1.97) | 0.018 |
|  |  | *P for trend* |  | 1.17 (1.07 ~ 1.27) | **<0.001** | 1.12 (1.03 ~ 1.22) | **0.012** | 1.07 (0.97 ~ 1.18) | 0.177 |
|  | Pre-DM | CTI (per 1 unit) | 500 | 1.05 (0.96 ~ 1.14) | 0.288 | 1.03 (0.95 ~ 1.13) | 0.470 | 0.97 (0.88 ~ 1.07) | 0.545 |
|  |  | Q1 | 68 | 1.00 (Reference) |  | 1.00 (Reference) |  | 1.00 (Reference) |  |
|  |  | Q2 | 116 | 1.25 (0.93 ~ 1.69) | 0.137 | 1.23 (0.91 ~ 1.66) | 0.176 | 1.03 (0.75 ~ 1.43) | 0.834 |
|  |  | Q3 | 170 | 1.54 (1.16 ~ 2.04) | **0.003** | 1.49 (1.12 ~ 1.97) | **0.006** | 1.33 (0.98 ~ 1.79) | 0.064 |
|  |  |  |  | HR (95%CI) | *p* | HR (95%CI) | *p* | HR (95%CI) | *p* |
|  |  | Q4 | 146 | 1.38 (1.04 ~ 1.85) | 0.027 | 1.31 (0.98 ~ 1.75) | 0.068 | 1.02 (0.75 ~ 1.40) | 0.890 |
|  |  | *P for trend* |  | 1.11 (1.02 ~ 1.20) | **0.016** | 1.09 (1.01 ~ 1.18) | 0.050 | 1.02 (0.93 ~ 1.12) | 0.638 |
|  | DM | CTI (per 1 unit) | 186 | 1.14 (1.00 ~ 1.32) | 0.058 | 1.11 (0.96 ~ 1.28) | 0.163 | 1.09 (0.92 ~ 1.28) | 0.312 |
|  |  | Q1 | 12 | 1.00 (Reference) |  | 1.00 (Reference) |  | 1.00 (Reference) |  |
|  |  | Q2 | 14 | 0.63 (0.29 ~ 1.37) | 0.245 | 0.62 (0.28 ~ 1.35) | 0.228 | 0.55 (0.22 ~ 1.42) | 0.218 |
|  |  | Q3 | 34 | 0.89 (0.46 ~ 1.72) | 0.730 | 0.83 (0.43 ~ 1.63) | 0.593 | 0.95 (0.44 ~ 2.03) | 0.885 |
|  |  | Q4 | 126 | 1.34 (0.74 ~ 2.43) | 0.328 | 1.23 (0.67 ~ 2.26) | 0.499 | 1.22 (0.60 ~ 2.47) | 0.579 |
|  |  | *P for trend* |  | 1.26 (1.06 ~ 1.50) | **0.008** | 1.23 (1.03 ~ 1.46) | **0.022** | 1.22 (0.99 ~ 1.49) | 0.058 |

HR: Hazard Ratio, CI: Confidence Interval.
Model 1: unadjusted for any covariates.
Model 2: adjusted for age, residence, marital status, education level, smoking status, and drinking status.
Model 3: adjusted for age, residence, marital status, education level, smoking status, drinking status, hypertension, dyslipidemia, SBP, DBP, and BMI.

In middle-aged participants, CTI was significantly associated with CVD risk in those with NGR (HR 1.57) but not in those with Pre-DM or DM. In elderly participants with Pre-DM, a higher CTI was linked to an increased risk of CVD, particularly in Q3 (HR 1.56), compared to Q1. No major differences were found in elderly individuals with NGR and DM (all *p* > 0.05). (Table 7)

**Table 7.**
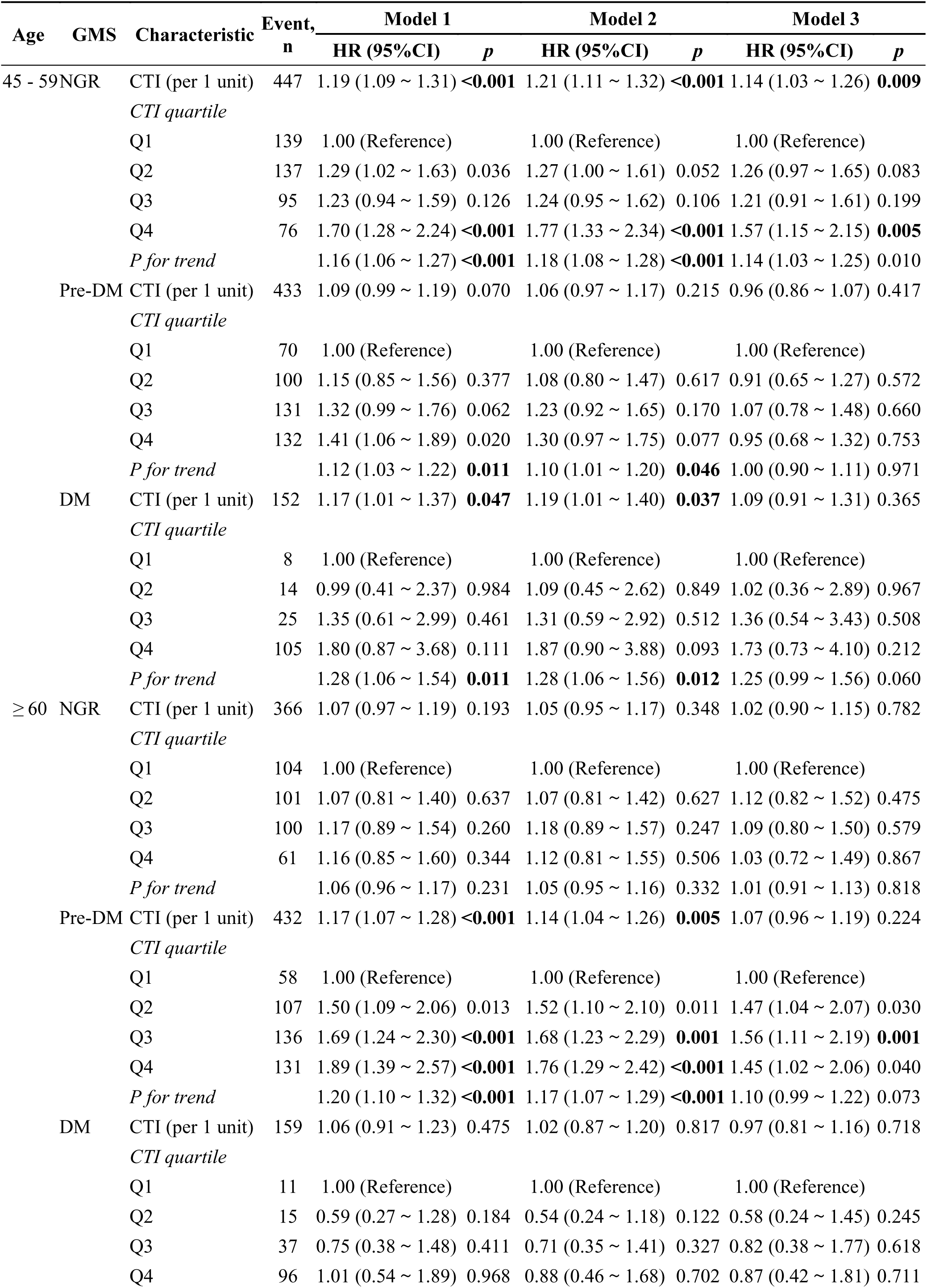
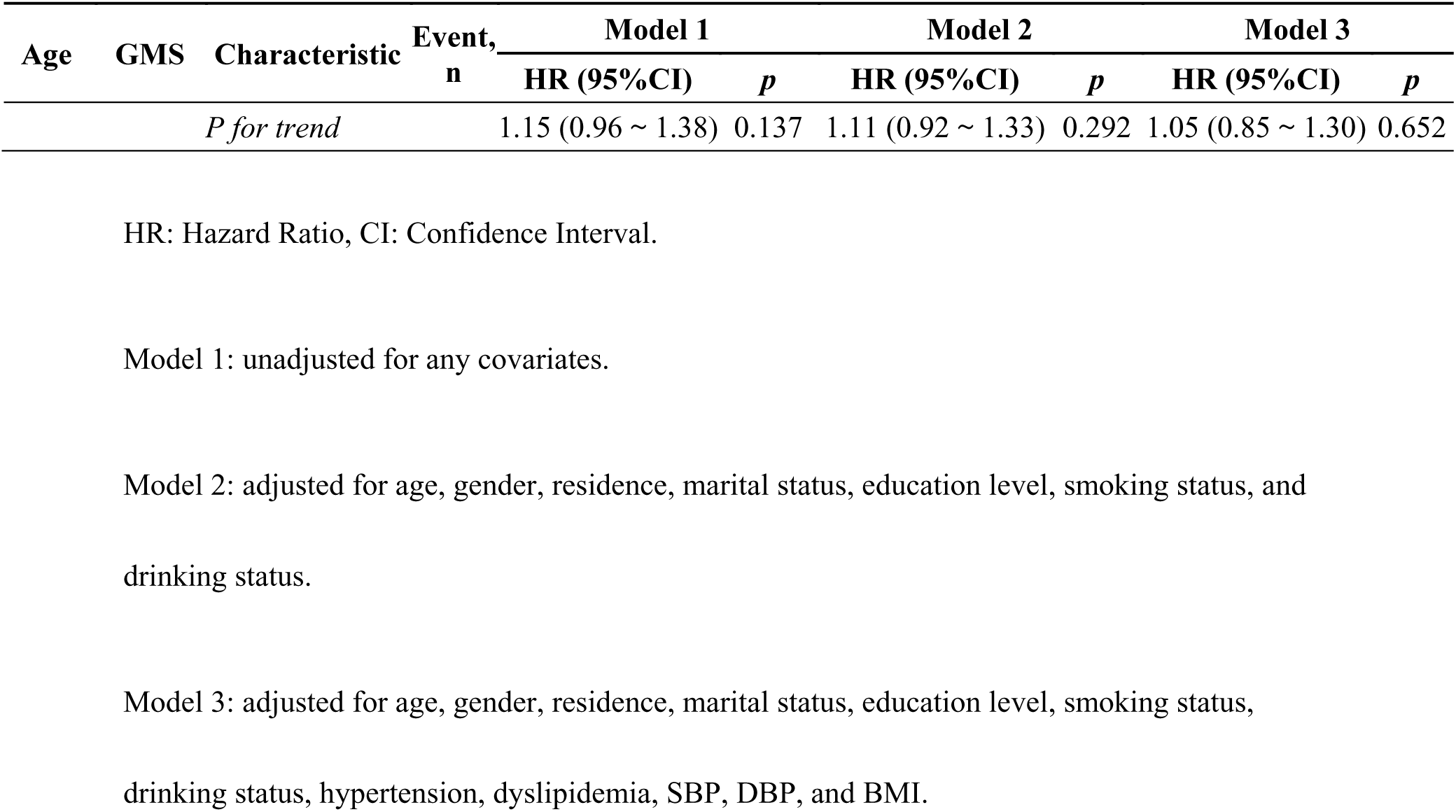
Association between the CTI and CVD incidence according to age and glucose regulation state.

### Subgroup analysis

Subgroup analysis showed consistent elevated CTI levels across various groups, including gender, age, smoking and drinking status, hypertension, dyslipidemia, glycemic statuses, and BMI. Without adjusting for covariates, CTI does not interact with any stratified variables (Fig. S4). Following adjustment, a notable interaction was identified between CTI and BMI (*p* = 0.042) (Fig. 6), but RCS analysis showed no significant CVD risk related to CTI in different BMI categories (Fig. S2 and S3). Notably, the correlation between CTI and CVD incidence was positive for BMI values less than 24, but no obvious relationship was observed for BMI groups of 24 to 27.9 and 28 and above (Table S5).

**Fig. 6.**
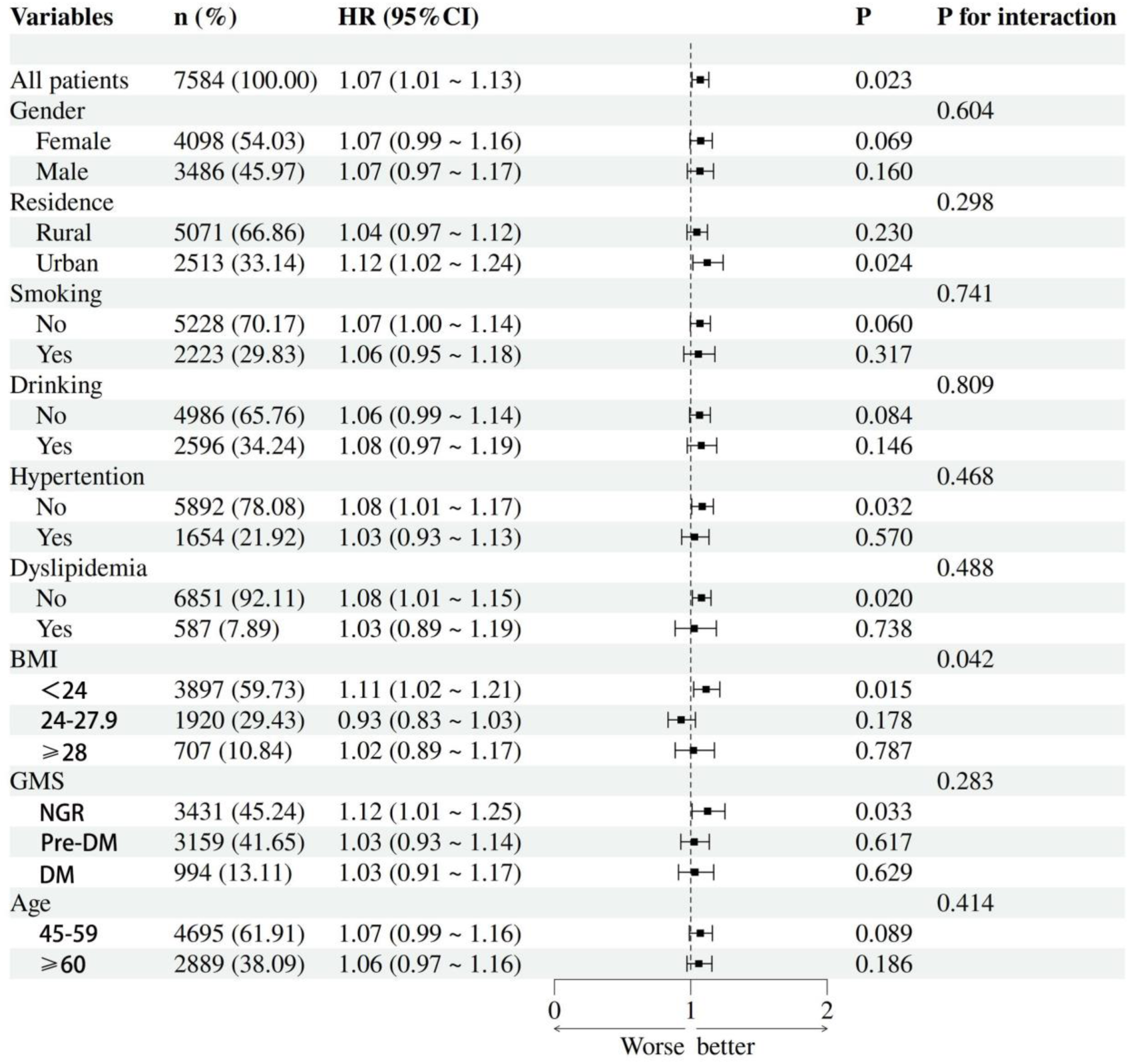
Subgroup and interaction analyses of the association between CTI and CVD risk. Adjusted for gender, age, residence, marital status, education level, smoking status, drinking status, hypertension, dyslipidemia, SBP, DBP, and BMI

### Sensitivity analysis

To ensure the findings’ robustness, we conducted several sensitivity analyses. Removing non-fasting participants (n=732) did not change the conclusions (Tables S6 - S9). Similarly, multiple interpolation and logistic regression models showed consistent results (Tables S10 - S13, S14 - S17). Additionally, the E-value for CTI from Model 3 was 1.31, indicating that significant unmeasured confounding could explain the observed hazard ratios.

## Discussion

This large-scale study is the first to identify a significant link between CTI and CVD risk, examining this relationship by gender, age, and glycemic status. It found a positive linear relationship between CTI and CVD occurrence, regardless of whether CTI was treated as a continuous or categorical variable. The association was consistent across genders and age groups, but varied by glycemic status: high CTI levels were linked to increased CVD risk in individuals with NGR and Pre-DM, but not in those with DM.

CTI, integrating markers of inflammation and IR, may serve as a biomarker for assessing CVD risk. Research indicates a positive linear relationship between CTI and CHD in the general American population, suggesting it could enhance CHD detection alongside traditional risk scores like the Framingham score^[27]^. CTI may also improve cardiovascular risk assessment and early CHD detection. Besides, higher CTI levels are linked to increased stroke risk in hypertensive patients, making it a potential stroke risk predictor^[35]^. The association between CTI and stroke is consistent across genders and age groups, and significant in individuals with normal glucose regulation and pre-diabetes^[26]^. In addition, elevated CTI levels are also linked to a higher risk of cardiometabolic multimorbidity, including diabetes, hypertension, and heart disease^[36]^. CTI, combining CTI with other indices, may offer a more comprehensive cardiometabolic risk assessment, indicating CTI’s value in preventing cardiometabolic diseases^[35]^. Consequently, the research indicates that CTI could be a dependable biomarker for stratifying CVD risk. For middle-aged and elderly people, CTI could be an effective biomarker for forecasting cardiovascular risk and predicting stroke, CHD, cardiometabolic diseases, and multimorbidity. More research is needed to confirm its broader applicability.

On the one hand, the TyG Index serves as a reliable surrogate marker for IR, showing high sensitivity and specificity to effectively predict disease risks^[36, 37]^. The TyG index showed a positive association with a higher rate of chest pain and was connected to all-cause mortality among those experiencing chest pain^[38]^. An initially high TyG index and its long-term trends were linked to hypertension risk, and recognizing a rising TyG index early might help in preventing hypertension in the future^[39]^. Higher and poorly managed TyG index levels were linked to a greater stroke risk^[40]^. A higher TyG index is an independent and causal risk factor for incident HF in the general population^[41]^. Non-diabetic young individuals with increased TyG index values have a higher probability of impaired cardiovascular fitness^[42]^. TyG–related parameters are useful for the early detection of non-alcoholic fatty liver disease (NAFLD) and metabolic-associated fatty liver disease (MAFLD)^[43]^. In conclusion, the TyG index, as a readily available biomarker, demonstrates broad clinical application potential. Its predictive value in cardiovascular diseases, cerebrovascular disorders, and metabolic-related conditions has been validated by multiple studies^[54–56]^. Nevertheless, a non-linear negative correlation between TyG and the conversion of glucose status from prediabetes to normoglycemia^[44]^. Physical impairments are greatly influenced by IR and prediabetes^[45]^. The use of the TyG index as a biomarker in CVD patients can be compromised by diabetes and high cholesterol^[46]^. Thus, different glucose conditions impact the link between TyG and cardiovascular risk. Despite TyG’s outstanding performance in risk assessment across various diseases, further research and validation are required to address applicability and standardization issues across different populations, thereby better guiding clinical practice. On the other hand, inflammation is another important risk factor for CVD^[47]^. Prior finding reveals that the risk of cardiovascular events rises with elevated serum CRP levels^[48]^. In CVD patients, CRP independently predicts heart failure^[49]^, and a causal link between CRP levels and stroke has been established through Mendelian randomization^[50]^. Cui et al.’s research highlights the combined impact of the TyG index and CRP on cardiovascular diseases^[51]^. Our study shows that the CTI consistently has a higher HR value than either CRP or the TyG index, whether considered as a continuous or categorical variable. ROC analysis suggests CTI has a better predictive value for CVD. This approach aids in identifying high-risk patients, allowing for precise risk assessments and targeted interventions. The superiority of the combined index (CTI) over the TyG index or CRP alone can be justified in various ways^[26]^. Initially, CTI gives a more detailed overview of an individual’s health, as the TyG index signals possible IR risk, while CRP sheds light on inflammation. The interaction of the two can uncover the joint influence of the “metabolism-inflammation” axis on CVD. Moreover, using only TyG or CRP focuses on a single aspect of possible pathological changes, while examining multiple indicators together can reveal variations across different metrics. Moreover, biomarker levels vary among individuals, with TyG and CRP providing different perspectives on IR and inflammation. This integrated method improves the capacity to detect and categorize individuals at greater risk.

Our study of 7,584 CHARLS participants found a significant link between higher CTI levels and CVD prevalence. RCS analysis showed a positive linear relationship between CTI levels and CVD events. Clinicians are advised to lower CTI levels to reduce CVD incidence in middle-aged and elderly individuals. Specifically, the relationship between CTI and CVD risk was alike for both genders, but the HR was elevated in males. A significant cohort study discovered that men have a much higher chance of both incident cardiovascular disease and cardiovascular mortality than women^[52]^. The lifetime risk of cardiovascular disease is higher in men than in women^[53]^. According to prospective cohort studies, men have roughly twice the lifetime risk of suffering from a myocardial infarction as women do, even when adjusting for standard atherosclerotic cardiovascular disease risk factors^[54]^. Earlier studies have indicated that men typically have a higher risk of stroke compared to women^[54]^. In essence, the differences in CVD risk between men and women are multifaceted, shaped by biological, hormonal, and socio-cultural factors^[56, 57]^. Furthermore, since middle-aged people show higher CVD risk than older adults, it is essential to concentrate on the CTI levels in this age group. Middle-aged individuals face a higher risk of cardiovascular and cerebrovascular issues than the elderly because they often maintain several unhealthy lifestyle habits, such as smoking, drinking alcohol, unhealthy diet, obesity, and insufficient physical activity^[7]^. Compared to the elderly, middle-aged people are more affected by cardiovascular risk factors like hypertension and dyslipidemia^[58]^. Thus, it is crucial to preserve healthy lifestyle habits from middle age and control cardiovascular risk factors to reduce the risk of cardiovascular and cerebrovascular diseases. We extended our analysis to stratify CTI and CVD risk by glycemic status, finding a significant non-linear positive correlation between CTI and CVD risk among individuals with NGR and Pre-DM. A retrospective study of over 260,000 participants without diabetes or CVD at the outset, the progression of FPG from normal to impaired and finally to a type 2 DM diagnosis was associated with a rising risk of heart attack, cardiovascular disease, and overall mortality^[59]^. According to a meta-analyses, non-diabetic people with an A1c level over 6.5% face twice the risk of cardiovascular death compared to those without diabetes and with A1c levels ranging from 5.0% to 6.0%^[60]^. In both the general population and those with atherosclerotic cardiovascular disease, Pre-DM was linked to a higher risk of all-cause mortality and cardiovascular disease^[61]^. The risk of CVD is elevated in IGT due to initial endothelial dysfunction and ongoing low-grade inflammation^[62]^. The findings imply that healthcare providers should aim to lower CTI levels in individuals with NGR and Pre-DM to lessen the risk of CVD. Howbeit, the connection was absent in those with DM, possibly explained by multiple factors. Initially, those with DM are persistently experiencing hyperglycemia. Their vascular endothelial injury, oxidative stress, and inflammation may have already reached a “plateau”, reducing the additional predictive benefit of CTI^[26]^. However, the metabolic abnormalities observed in non-diabetic individuals (NGR, Pre-DM) are relatively mild, and CTI may serve as a more sensitive indicator for assessing the risk of early vascular injury.

Secondly, IR exhibits a "threshold effect"^[63]^. In non-diabetic individuals, the extent of IR is positively correlated with elevated CTI levels. Conversely, in DM patients, IR generally has already initiated significant activation of downstream signaling pathways. Further increases in CTI do not correspond to a heightened risk. Besides, individuals with DM often have other conditions like hypertension and dyslipidemia, which can complicate the direct link between CTI and CVD. In individuals without DM, metabolic disturbances are the key factors in CVD risk, pointing to the predictive value of CTI for this population. Moreover, CTI may fail to account for personal differences among DM patients, causing a greater CVD prediction in NGR and Pre-DM groups than in DM groups^[64]^. Furthermore, the metabolic issues linked to DM might increase CVD risk via different mechanisms, like microvascular dysfunction, obesity, and hypertension, etc., possibly reaching a saturation level where CTI ceases to offer further predictive insights^[65]^. Additionally, DM is indicated by the pancreas excessively secreting the peptide amylin, which aggregates into toxic deposits in vessel walls, leading to microvascular issues^[66]^. Last but not the least, blood glucose regulation varies greatly among people with diabetes. The pathological features of poorly managed diabetes can vary greatly from those of well-managed diabetes. Thus, this research highlights the significance of adjusting CTI levels in patients based on their glucose metabolism states.

We analyzed the link between CTI and CVD across gender and age under different glycemic statuses. The data reveal that the link between CTI and CVD risk is similarly patterned in males with Pre-DM and females with NGR. This implies that keeping CTI levels low is vital for males with Pre-DM and females with NGR. Additional analysis was performed on middle-aged and older adults with varying glycemic conditions. The findings indicated that for middle-aged individuals, CTI is strongly linked to a higher risk of CVD in those with NGR. With a notable CVD risk increase in middle-aged NGR patients in Q4 versus Q1. this highlights the importance of monitoring CTI in middle-aged NGR patients to prevent CVD. Hence, it is necessary to reduce CTI levels to prevent CVD in this demographic. However, in stratified analyses by gender, age, and glycemic status, the RCS did not show a significant correlation, unlike the Cox regression analysis. This discrepancy may arise from methodological differences, population heterogeneity, and limited statistical power due to small subgroup sizes. Subgroup analyses highlighted an interaction between CTI and BMI levels. RCS analysis found no significant link between CTI and CVD risk across BMI levels, while Cox regression showed a significant positive correlation only for BMI < 24. Therefore, reducing CTI levels is recommended for individuals with BMI < 24. This study is, as far as we are aware, a novel examination of the relationship between CTI and CVD risk based on gender, age, and glycemic status.

The exact link between CTI and CVD risk is unclear, but several factors may explain it. IR and inflammation can cause endothelial dysfunction, reduce nitric oxide use, disrupt coagulation, and speed up atherosclerosis, raising CVD risk^[67, 68]^. Metabolic and vascular IR together heighten CVD risk in diabetes and related conditions^[69]^. Inflammation worsens IR, releasing inflammatory mediators and enhancing systemic inflammation^[70]^. This mutual aggravation of inflammation and IR increases CVD risk^[71]^. Lipid abnormalities and endothelial dysfunction in insulin-resistant states contribute to atherosclerotic plaque formation^[72]^. Additionally, inflammation and IR destabilize these plaques, increasing rupture risk, which can lead to thrombosis and CVD^[73]^. Patients with IR and inflammation often have comorbidities^[74]^ like hypertension^[75]^, diabetes^[76]^, and obesity^[77]^, which are major CVD risk factors. Higher CTI levels may indicate more severe vascular injury and increased CVD incidence, though the exact mechanism is unclear and requires further study.

This research highlights CTI’s role in CVD across different demographics, highlighting the importance of tailored risk management strategies based on gender, age, and glycemic status to prevent cardiovascular disease. With rising global CVD cases, especially among those with inflammation and IR, timely identification of high-risk patients is crucial. To calculate CTI, only standard fasting blood tests are needed, which include CRP, TG, and blood glucose measurements. This method is economical and ideal for screening large populations. Our findings can help clinicians manage CTI levels to prevent CVD individually.

### Strengths and limitations

This research offers several advantages: it is the first to examine the link between CTI and CVD incidence by gender, age, and glycemic status, emphasizing population heterogeneity; it uses a prospective, nationwide longitudinal cohort design with a large sample size, enhancing reliability; it includes subgroup and sensitivity analyses for consistent results across different populations, aiding clinical practice; the longitudinal design of CHARLS reduces the possibility of reverse causality; and for practical purposes, CTI is easily accessible and can identify high-risk individuals that traditional risk assessments miss.

However, several limitations of this study should be recognized. Firstly, like any observational study, causality cannot be confirmed between CTI and CVD. To substantiate the evidence, further genetic studies and clinical trials are essential. Besides, heart disease and stroke diagnoses are self-reported, which might underestimate true CVD incidence, though prior validation supports their reliability^[78, 79]^. Thirdly, the CHARLS questionnaire does not classify heart disease and stroke subtypes, limiting specific disease risk analysis. Furthermore, despite multivariate adjustments, unknown confounders (such as dietary habits, fitness routines, hereditary traits, or pharmaceutical intake) might still affect the results. The observed relationship could be inflated if these factors are related to both CTI and cardiovascular risk, though the E-value suggests our findings are robust. Moreover, some analyses are based on baseline data (e.g., CRP, TyG, and glycemic status), which may disappreciate the dynamic impact of long-term exposure on cardiovascular events due to potential survival bias. While longitudinal follow-up data have somewhat addressed this problem, it’s important to recognize that initial measurements might not completely represent the long-term metabolic condition. Future research should address to better understand its relationship with CTI levels and CVD. Additionally, the study focuses only on middle-aged and elderly individuals in China, potentially limiting its applicability to other populations. Future research should validate CTI’s predictive effect on stroke in diverse groups and settings, and intervention studies could explore whether modifying CTI reduces CVD risk.

## Conclusion

The study found a strong positive link between CTI and CVD risk, consistent across genders and age groups. High CTI levels increased CVD risk in those with NGR and Pre-DM, but not in those with DM. These findings highlight the importance of tailored risk management strategies based on gender, age, and glycemic status to prevent CVD.

## Data Availability

The data supporting the findings of this study are available on the CHARLS website (http://charls.pku.edu.cn/).

## Acknowledgements

This study utilized data from the CHARLS database. The authors express their gratitude to the CHARLS research team and all individuals who participated in the study.

## Author contributions

S.Q., C.Y. and S.G. conceived and designed the study, curated the data, and wrote the main manuscript text. J.X., J.W. and X.L. directed the conception of the manuscript and performed medical review of the analysis plan and first interpretable results. X.W.L. and Q.X.Z. carried out statistical analysis. H.Z., K.C. and Y.W. prepared methodology and figures. G.C., Y.C., and Q.C.Z. conducted the literature search and performed critical revision. All authors reviewed and approved the final manuscript.

## Declarations

### Ethics approval and consent to participate

CHARLS was approved by the Institutional Review Board of Peking University (approval number: IRB00001052-11015 for the household survey and IRB00001052-11014 for blood samples), and all participants provided written informed consent.

